# Social Distancing is Effective at Mitigating COVID-19 Transmission in the United States

**DOI:** 10.1101/2020.05.07.20092353

**Authors:** Hamada S. Badr, Hongru Du, Max Marshall, Ensheng Dong, Marietta Squire, Lauren M. Gardner

## Abstract

COVID-19 is present in every state and over 90 percent of all counties in the United States. Decentralized government efforts to reduce spread, combined with the complex dynamics of human mobility and the variable intensity of local outbreaks makes assessing the effect of large-scale social distancing on COVID-19 transmission in the U.S.a challenge. We generate a novel metric to represent social distancing behavior derived from mobile phone data and examine its relationship with COVID-19 case reports at the county level. Our analysis reveals that social distancing is strongly correlated with decreased COVID-19 case growth rates for the 25 most affected counties in the United States, with a lag period consistent with the incubation time of SARS-CoV-2. We also demonstrate evidence that social distancing was already under way in many U.S. counties before state or local-level policies were implemented. This study strongly supports social distancing as an effective way to mitigate COVID-19 transmission in the United States.

## One Sentence Summary

Social distancing within the United States is slowing the spread of COVID-19, with a lagged effect of nine to twelve days.

A cluster of cases of pneumonia of unknown cause in Wuhan, China was first reported on December 31, 2019 (*1*), and a week later identified as a novel coronavirus, COVID-19 (*2*). COVID-19 has since spread rapidly around the world, nearing 4 million confirmed cases and over 250,000 deaths reported in 187 countries/regions as of May 5 (*3*). The first case of COVID-19 in the U.S. was reported on January 20 in Snohomish County Washington (*4*), and as of May 5, COVID-19 has been reported in every U.S. state and over 2800 U.S. counties. (*5*). Until the widespread availability of a vaccine, social distancing will remain one of the primary control mechanisms for mitigating the spread of COVID-19.

In China, a nationally coordinated effort limiting travel and social interaction effectively mitigated the spread of the disease (*6*). Critically, in contrast to the nationally mandated directives put in place in China, the U.S. directives to “shelter in place” and temporarily close non-essential businesses and schools were made at the state and local level throughout March and April 2020 (Fig. S1 and Table S1). This distributed decision-making process and enforcement has resulted in an outbreak mitigation response that is highly variable in both space and time. Adding to this complexity is the varying intensities of the outbreak around the U.S., with some counties nearing their peak while others remain in the early stages of an epidemic (*5*). Together, these issues pose a significant challenge to evaluating the effectiveness of social-distancing policies in the U.S. To address this issue, we use real-time mobility data derived from mobile phones to quantify the progression of social distancing within the U.S. Subsequently, we examine its relationship to the rate of emerging COVID-19 cases in 25 U.S. counties with the highest number of reported cases as of April 16, 2020 (Fig. S2, Table S2). Our analysis provides strong evidence that social distancing is leading to a decrease in the rate of new cases in these counties, and is therefore an effective mitigation policy for COVID-19 in the U.S.

Previous studies have evaluated the connection between travel and transmission of COVID-19, but they are restricted to examining the disease in China. In addition to the aforementioned work by Kraemer et al. (*6*), Zhao et al. (2020) found a positive association between confirmed cases and the quantity of domestic passenger travel within 10 cities outside of Hubei Province (*7*). Tian et al. (2020) found evidence that social distancing measures in cities throughout China, delayed case transmission (*8*). Chinazzi et al. 2020 (*9*) used a transmission model to project the impact of travel limitations on the spread of COVID-19 in China, finding travel restrictions to the affected areas to have modest effects and that transmission reduction interventions are more effective at mitigating the pandemic. These studies are encouraging and suggest social distancing measures should successfully mitigate infection transmission outside of China, however this has yet to be shown. There are qualitative studies and projections for social distancing helping to reduce the spread of COVID in countries such as Italy (*10*) and the US (*11*) but, to date, no such quantitative analysis has been conducted outside of China. We therefore extend the current body of work to evaluate the impact of social distancing on the spread of COVID-19 in the United States, the country which has reported the most confirmed cases and deaths due to COVID-19 in the world.

### Social Distancing is occurring in the U.S

To quantify the amount of social distancing in each U.S. county, we define a social distancing ratio (SD) for each day (t) and county (j). Our social distancing ratio, SD, reflects the relative change in the number of individual trips made in each county, each day, relative to ordinary behavioral patterns (prior to COVID-19). To compute this measure, we use daily origin-destination (OD) trip matrices at the U.S. county level derived from mobile phone records obtained from Teralytics (*12*). This effort aligns with a recent Letter in Science (*13*) supporting the use of aggregated mobility data to monitor the effectiveness of social distancing interventions. The data provided consists of the number of unique daily trips made between all pairs of US counties, each day, from January 1 through April 20, 2020. Specifically, SD is the sum of the total trips incoming, outgoing and within each county on a given day, divided by the same measure on a baseline day. The baseline value is specific to each day of the week and taken as the average over the last three weeks in January 2020, when travel patterns were stable (see Fig. S3). We interpret this metric as a proxy for social distancing based on the assumption that when individuals make fewer trips, they physically interact less. Formally, 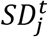 is calculated as follows:

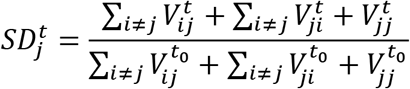

Where 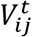 represents the number of trips from county *i* to *j* on day *t*, and *t*_0_ represents the baseline measure. This metric accounts for movements both between and within counties, thus includes changes in typical commuting patterns as well as micro-level (within county) movements, (e.g., travel to local grocery stores, shopping centers, gyms, schools, etc.) Using this function, an SD of zero would indicate no trips were made, while a value of 0.5 indicates half the number of trips relative to the baseline were made on a given day. An SD of one signifies no change in behavior since the advent of COVID-19 in the US, and any value above one means that mobility has actually increased from the baseline.

The differences in county-level SD across the U.S. from January 24 to April 17, 2020, are illustrated in Fig. 1 and shown at a daily resolution in the Animation S1. For the 25 U.S. counties with the highest number of reported cases, the SD as of April 17 are shown in Fig. S4, with the 25 counties ranked accordingly. The SD for these 25 counties ranges from 0.35 in New York City, to 0.63 in Harris County, Texas, which illustrates varying SD measures and associated behavioral change in place. Counties illustrating the most social distancing in the first week of April are predominantly in New York, New Jersey, and Massachusetts, the locations reporting the most COVID-19 cases to date.

**Fig. 1.**
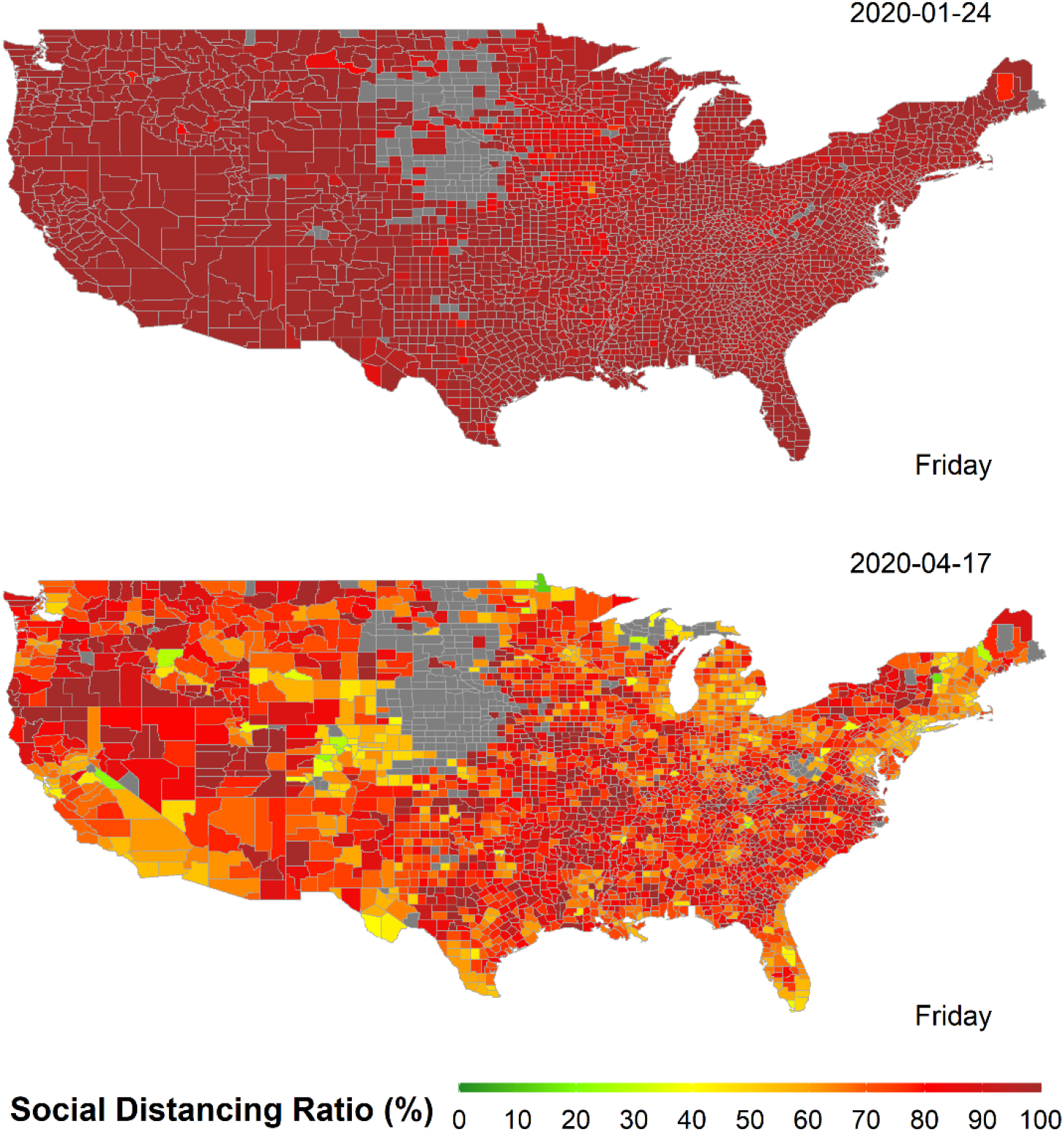
Social distancing ratio (SD) per US county on Friday, January 24, 2020 (left), and on Friday, April 17, 2020 (right). The greyed-out area in the Midwest are filtered due to low coverage in the Teralytics data set (*12*). This includes all counties with a total trip counts less than two standard deviations bellow the mean.

For the set of 11 states corresponding to the top 25 counties (shown in Fig. S5), similar SD behavior is observed at the state level. Fig. S6 provides the complete list of all U.S. states ranked by their respective SD ratios as of April 17. Consistent with county level behavior, there is evidence of different social distancing across states, with only D.C. reducing trips below 50%, and the rest of the 50 states moving around between 53% to 90% traditional levels. Many of the southern states, which implemented temporary closure of non-essential business later in March or early April, report higher SD ratios. Fig. 2 illustrates the state SD trends over time, in relation to the introduction of each state’s social distancing directive, which are noted by the red dashed vertical lines (explicit dates are listed in Table S1). In addition to the timing of the state-level directives, we collected information on the county level social distancing directives that were implemented in each of the 25 counties of focus. The timing of the local-level directives relative to SD behavior at the state level are also illustrated in Fig 2, as the blue dashed lines. A list of all local directives and respective dates is provided in Table S3 and includes the time gap between local and state-level actions. Fig 2 illustrates that social distancing began to occur in early March, well before the first U.S. state (California), implemented a “stay at home” directive, on March 19. The county level directives partially explain the earlier decline in SD, which is observed to begin well before the state-level directives where put in place, however all states illustrate some level of social distancing even before county level directives were enacted. Fig. S7 and S8 further illustrate the relationship between state and local directives and social distancing ratio (SD) for each of the selected 25 counties based on total (S7) and internal/local-only (S8) trips, respectively. From this county level break down it is evident that locations such Bergen, NJ, Oakland, MI, Orange, NY and Fairfield, CN implemented local-level directives much earlier than their corresponding states, which align with the start of social distancing decline in these counties. In contrast, in Orleans and Jefferson, LA where local level directives were implemented late or not at all, social distancing was declining for weeks without directives in place, suggesting other motivations drove this behavioral change within the region. For all 25 counties, except for Jefferson, LA, local directives were implemented at least three to 17 days prior to the state level stay-at-home directives, with an average difference of 7.3 days.

**Fig. 2.**
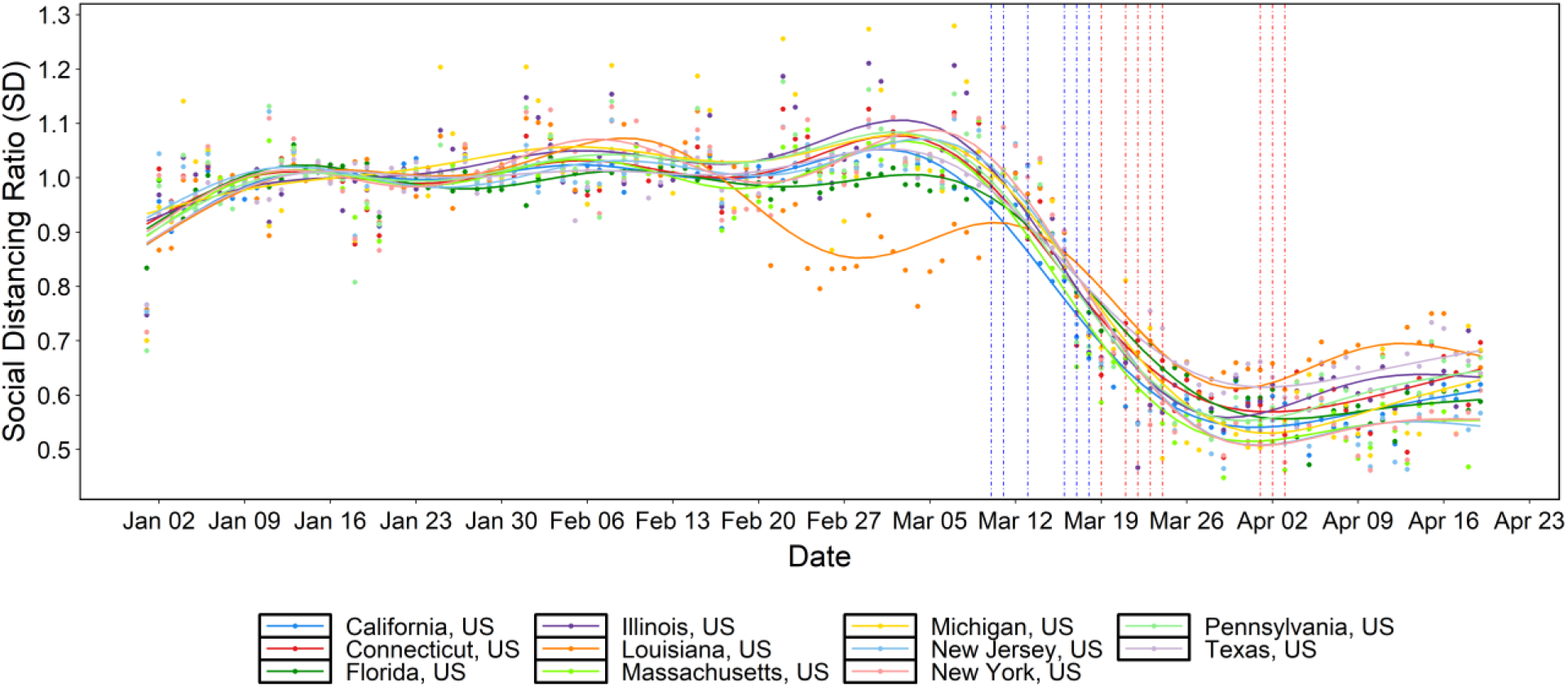
Social distancing ratio (SD) timeseries (relative to the last three weeks of January 2020) for US states and the corresponding dates of stay-at-home orders (vertical dashed red lines). Blue and red vertical dashed lines represent dates of implementation for local and state social distancing directives, respectively (some overlap). The dots represent the raw SD data while the plotted lines are smoothed using generalized additive model (GAM).

### COVID-19 Growth Rates are decreasing in some parts of the U.S

Epidemiological data from the JHU CSSE COVID-19 dashboard (*5*), which includes daily data on cases and deaths for each US county, is used to compute the COVID-19 growth rate ratio (GR) for a given county on a given day. The ratio is defined as the logarithmic rate of change (number of newly reported cases) over the previous three days relative to the logarithmic rate of change over the previous week. The growth rate ratio for any county *j* on a day *t* is calculated as follows:

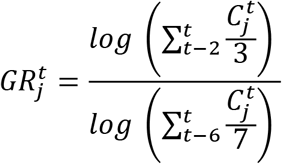

where 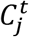 is the number of new cases reported in county *j* on day *t*.

GR can take on any non-negative value. A GR equal to zero indicates no new confirmed cases were reported in the last three days, while a value below one means that the growth rate during the last three days is lower than that of the last week. A value greater than one represents a growth rate increase in the last three days. We use 3-day moving averages to smooth volatile case reporting data. This metric, used in conjunction with the social distancing ratio SD, allows us to grasp the complex and time-dependent dynamics at play between human mobility and COVID-19 spread for each county in the US.

## Social distancing leads to decreasing COVID-19 transmission the U.S

Using the two metrics introduced above, we evaluate if and how well social distancing influences the rate of new infections in the twenty-five counties in the United States with the highest number of confirmed cases on April 16, 2020. King County, Washington is excluded because it precedes widespread social distancing and was driven by an infection source that differs from other outbreaks in the US. We fit a linear regression model for each county, specifically using lagged SD as a predictor of COVID-19 growth rate and test the correlation of SD and GR at different time lags. From these results, the correlation between the SD and GR is computed for each county. Additionally, the most highly correlated lag range is identified based on the (maximum) mean and (minimum) standard deviation of the correlations across all counties, based on Pearson correlation coefficient. The residuals confirm that SD and GR are linearly related.

An optimal lag of 11 days, with a window of 9 to 12 days (see Fig. 3) is identified from the county level analysis. This lag represents the period separating the beginning of social distancing and onset of case growth reduction. An interval of this length is consistent with the estimated four to five-day (median) incubation period of the virus (*14-16*). In other words, the lag time that best links social distancing and case growth rate in our analysis reflects the time it takes for symptoms to manifest after infection, worsen, and be reported. The confidence intervals shown on the curve illustrate that this estimated lag interval is robust and consistent across multiple counties.

**Fig. 3.**
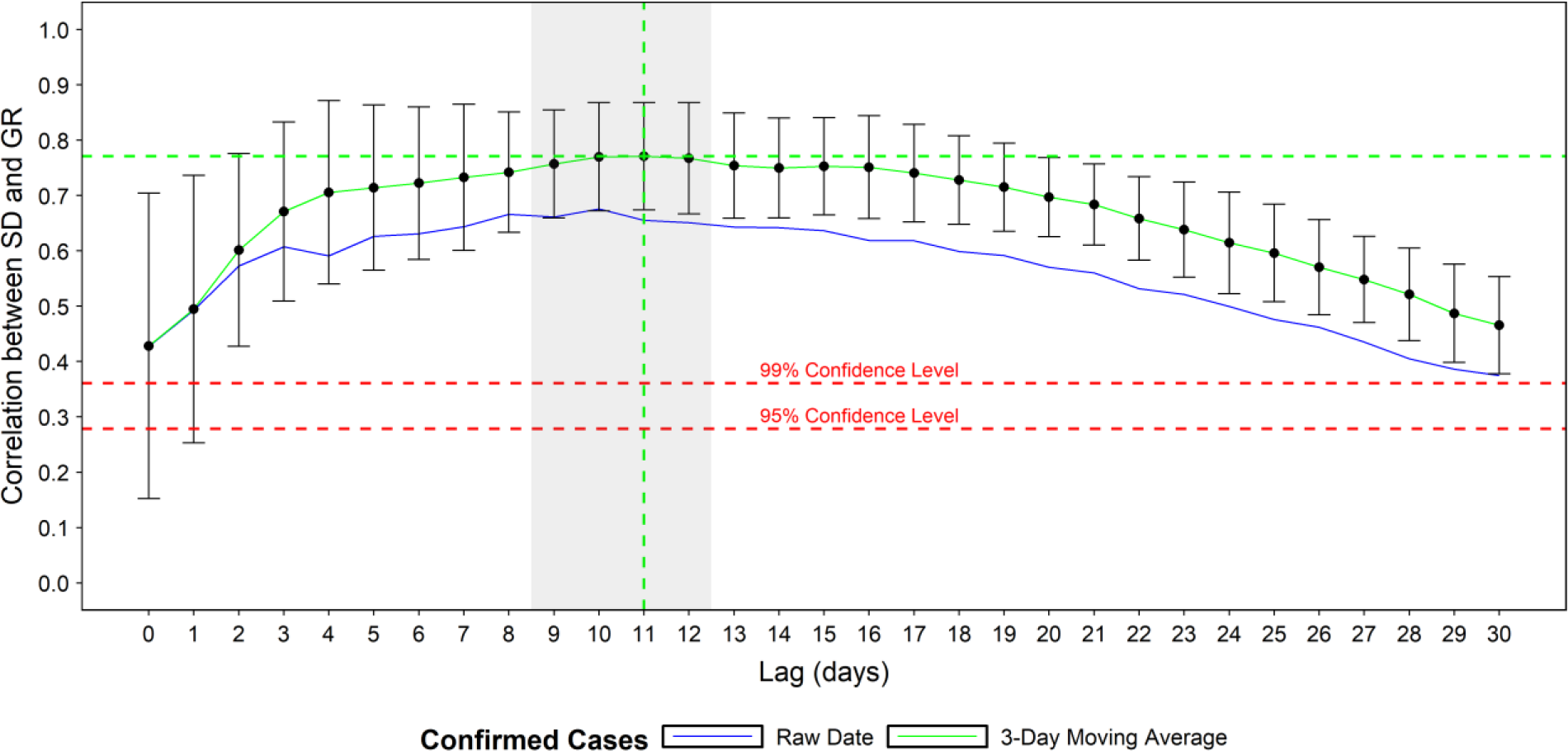
Mean and standard deviation of the county correlations between social distancing ratio and growth rate ratio at different lags (in days). Correlations found to be significant at a 95% confidence level.

Fig. 4 illustrates the county-specific correlations between SD and GR, for an eleven-day lag (A), alongside the (B) computed GR, (C) SD, and (D) number of daily COVID-19 cases spanning March 16 to May 4. The case data illustrates the number of new daily cases for all 25 counties increases through March, then slows in April in many of counties, and begins decreasing in some counties. During the same period, the social distancing ratio steadily decreases, specifically during the second half of March, before stabilizing for most locations in early April, followed by a slight increase throughout April.

As indicated by the red and blue dashed lines, which represent the timing of the state and county level social distancing directives, in almost all counties the initial decline in SD began prior to any formal regulation being put in place, and can therefore be attributed to proactive behavioral changes among the local population. The exact motivation behind the individual-level behavioral changes requires further study but could possibly be attributed to media and information sharing, which itself varies substantially by information source, location, and time. The growth rate ratio shows a general decreasing trend for all locations through March, stabilizing around one in April, which is consistent with the case curves. All correlations are found to be significant at a 95% confidence level, with the correlations for many of the top 25 counties above 80%. These high correlations suggest that social distancing has had a significant effect on the spread of COVID-19 in the U.S. Some exceptions include Orleans, LA (.61) and Harris County, TX (.53), the former of which can be attributed to an outbreak which took off before social distancing was common practice, while the case counts in Harris, TX are likely underestimated due to limited testing rates in Texas.

**Fig. 4.**
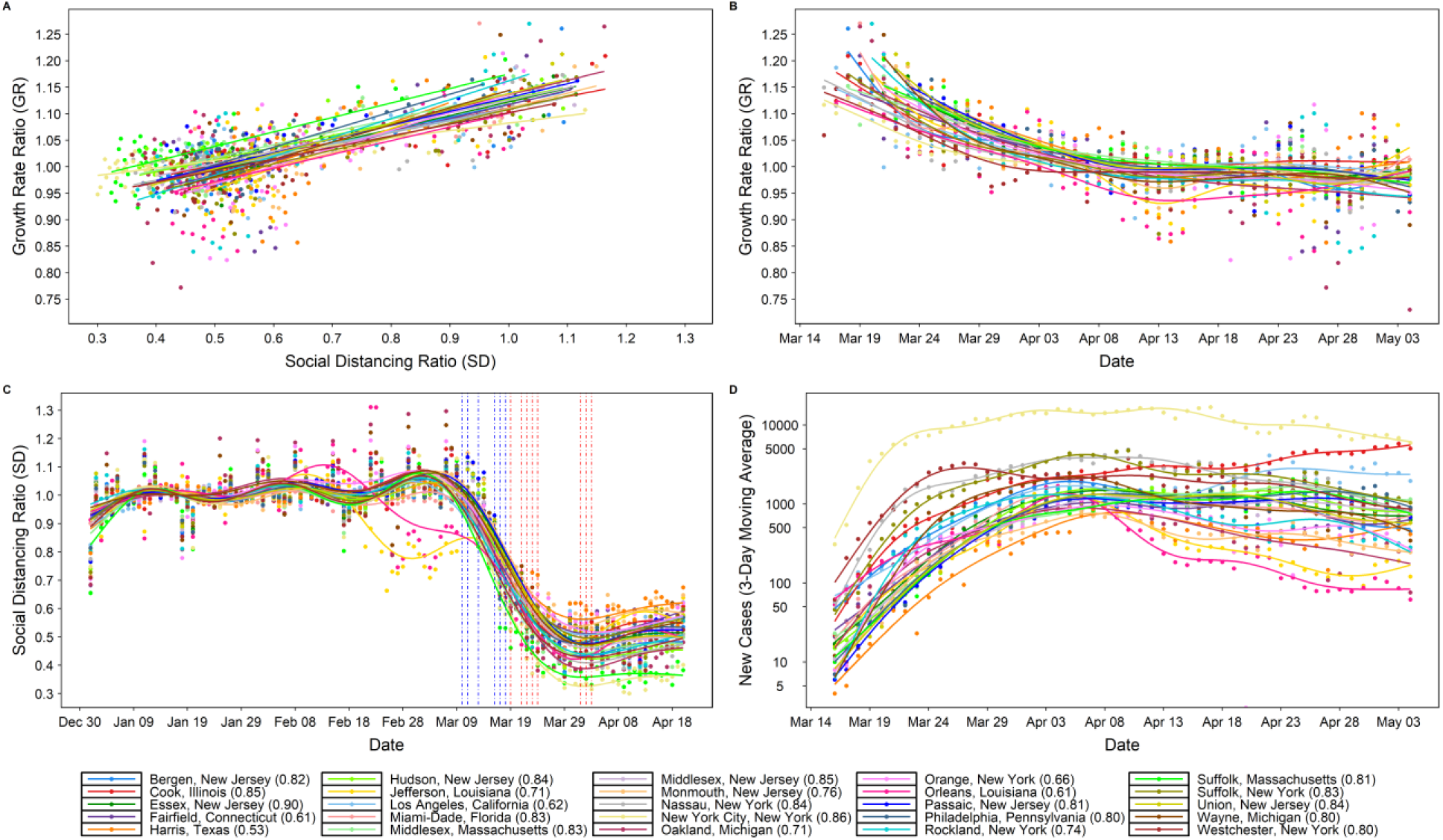
Relationship between social distancing ratio (SD) and growth rate ratio (GR) given a eleven-day lag (A), with (B) growth rate ratio GR over time (C) progression of social distancing ratio SD over time and (D) new confirmed cases over time. Correlations found to be significant at a 95% confidence level. In (C), the dates of state-level stay-at-home orders are shown as vertical dashed red lines, while the local-level social distancing orders are shown as dashed blue lines. The dots represent the raw data while the plotted lines are smoothed using generalized additive model (GAM).

We repeated the equivalent evaluation method at the state level, using aggregated case reports from all the counties within each state. The results are consistent with the county-level analysis, revealing a 9-12-day lag window (Fig. S9), and displaying similarly high correlations between social distancing and growth rate (Fig. S10). The lowest correlation, 0.69, is observed for Massachusetts, which is likely a result of intra-state noise in the data and spatial variability, specifically with regards to the state of the outbreak, testing capabilities, and varied degrees of behavioral change associated with social distancing measures.

## Discussion

The United States has enacted a complex combination of responses to COVID-19. Government policy varies in space, scale, and time (Table S2 and S3), resulting in varied patterns of movement and behavioral changes throughout the country. Simultaneously, the progression and intensity of local outbreaks differs markedly by location. This landscape makes quantifying the impact of social distancing on COVID-19 spread a nontrivial task. Nevertheless, our methodology captures relevant trends in human behavior as it relates to the spread of the disease. Because our analysis uses real-time mobility data at the individual level, we capture the dynamics of social distancing without relying on assumed efficacy of shelter-in-place orders. Additionally, we use the real-world frequency of trips, not extrapolated transmission rates or travel distances, as our mobility indicator. This means our social distancing metric is driven purely by how much people actually move, both between and within counties. Since our case data consists simply of reported cases at the county level, our analysis is a powerful comparison of actual human behavior and the documented reality of COVID-19 in the US.

Our results use this real-world data to show that social distancing is a useful technique to help the United States “flatten the curve” of new cases, demonstrating a strong and statistically significant correlation between social distancing and reduction of COVID-19 case growth. Importantly, our statistical analysis reveals that the effect of social distancing on decreasing transmission is not perceptible for nine to twelve days after implementation. Besides emphasizing its necessity, this study also reveals that social distancing (and outbreak growth deceleration) in the counties most affected by COVID was driven primarily by local-level regulations and changes in individual-level behavior; the state (and federal) actions implemented were done so either too late (or not at all). This is an important insight, as it demonstrates (given the clear correlation we present between social distancing and case growth), that it is within the power of each U.S. resident, even without government mandates, to help slow the spread of COVID-19. Critically, if individual-level and local actions were not taken, and social distancing behavior was delayed until the state-level directives were first implemented, COVID-19 would have been able to circulate unmitigated for additional weeks in most locations, inevitably resulting in more infections and lives lost. Further, the strong relationship between social distancing and outbreak case growth rates suggest that a return to ‘normal’ interaction patterns will result in an increase in case growth rates, which may appear 9-12 days after behavioral changes ensue. However, under these changes, additional precautions such as hand washing, wearing masks and self-isolation when sick may help to lessen the case growth rates.

This study is subject to multiple limitations. First, we focus on quantifying the relationship between social distancing and case growth rates, therefore the role of other potential mitigating factors (e.g., wearing masks, hand washing, etc.) that could also have contributed to the decline in the case growth rate observed during March are not accounted for. Second, we use the growth rate ratio (GR) as our representative variable for the degree of transmission occurring in a region. We believe this is an intuitive and representative estimate for the spread of COVID-19 amongst a local population, but future extensions of this analysis can explore replacing this variable with more traditional transmission indexes commonly used in infectious disease epidemiology. Third, the case data is error prone due to both reporting issues and limited testing capacity, especially in early March before widespread testing was underway. We partially address this issue by using a 3-day moving average for the case data. Forth, the analysis is focused on 25 counties which may represent a biased sample of locations; however, the same results are shown to hold when extrapolated up to the state level, which lends additional confidence to the conclusions. Last, the data used in this analysis does not differentiate amongst sociodemographic groups, and therefore may not representatively capture all groups such as the elderly, low income families and under-representative minorities, for whom social distancing may not be an option, or may not have cell phones.

In conclusion, our results strongly support the conclusion that social distancing pays dividends in the vital reduction of load on hospital systems in the United States. It may be difficult to recognize the value in safe behavior when the reward is not obvious, and the danger is not immediate. This is particularly true given the economic and social repercussions of the COVID-19 response. Nevertheless, given the lack of proven antiviral drugs or a vaccine, social distancing is the most important and timely way to combat the spread of COVID-19 (*17,18*). These findings also highlight the difference in pandemic control policy between the U.S. and China and should serve to support more timely policymaking in the U.S. moving forward. This is particularly relevant as the U.S. begins to loosen stay at home orders, once again doing so in a highly decentralized manner. We hope that our results will motivate both individuals and decision makers in the US to take seriously the importance of advocating for safe and data-driven policy in the face of this pandemic, while balancing its impact on associated communities.

## Data Availability

All data used for the analysis are provided as supplementary files

## Supplementary Materials for

**This PDF file includes:**

Supplementary Text

Figs. S1 to S10

Tables S1 to S3

Caption for Animation S1

Captions for Data S1 to S2

## Supplementary Text

### Mobility Data

The real-time mobility data used in this analysis, initially provided as daily origin-destination (OD) matrices, was provided by Teralytics (*12*). The data is based on anonymized mobile network data, covering 86 Million people with a good coverage across all demographics and geos (with the exception of some midwestern states). From each phone they receive on average 150 cell tower pings evenly spread over each day. Teralytics processes the raw network event data and turn this into trip data by determining moving and stationary activity. The trip data is extrapolated up to represent the entire population by determining the home area of each phone and then compute an extrapolation factor for each phone based on the number of phones observed to have home locations in the area relative to the census population of the area. Through measuring mobile phones, this data includes every movement across all modes, including plane, car, public transit and walking. For the state level analysis, we filter out those counties in states with low coverage, specifically those with trip counts less than two standard deviations bellow the mean. This filtering is shown in the greyed-out area in the Midwest in Animation S1. The 25 counties that are the focus of this analysis have good coverage. The data used in this analysis is provided as a supplementary file.

**Fig. 1.**
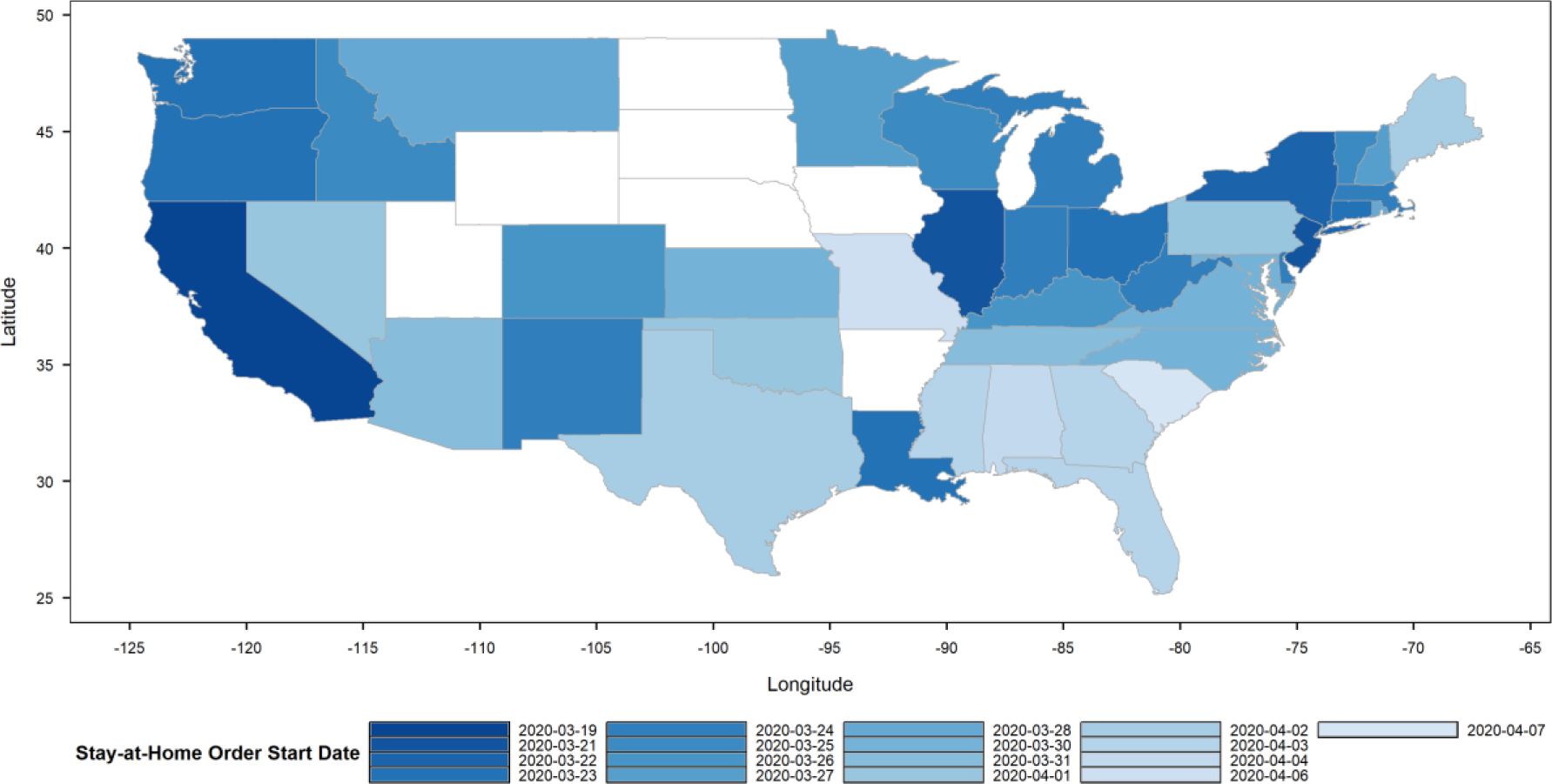
The start dates of stay-at-home orders for all U.S. states throughout March and April 2020.

**Fig. 2.**
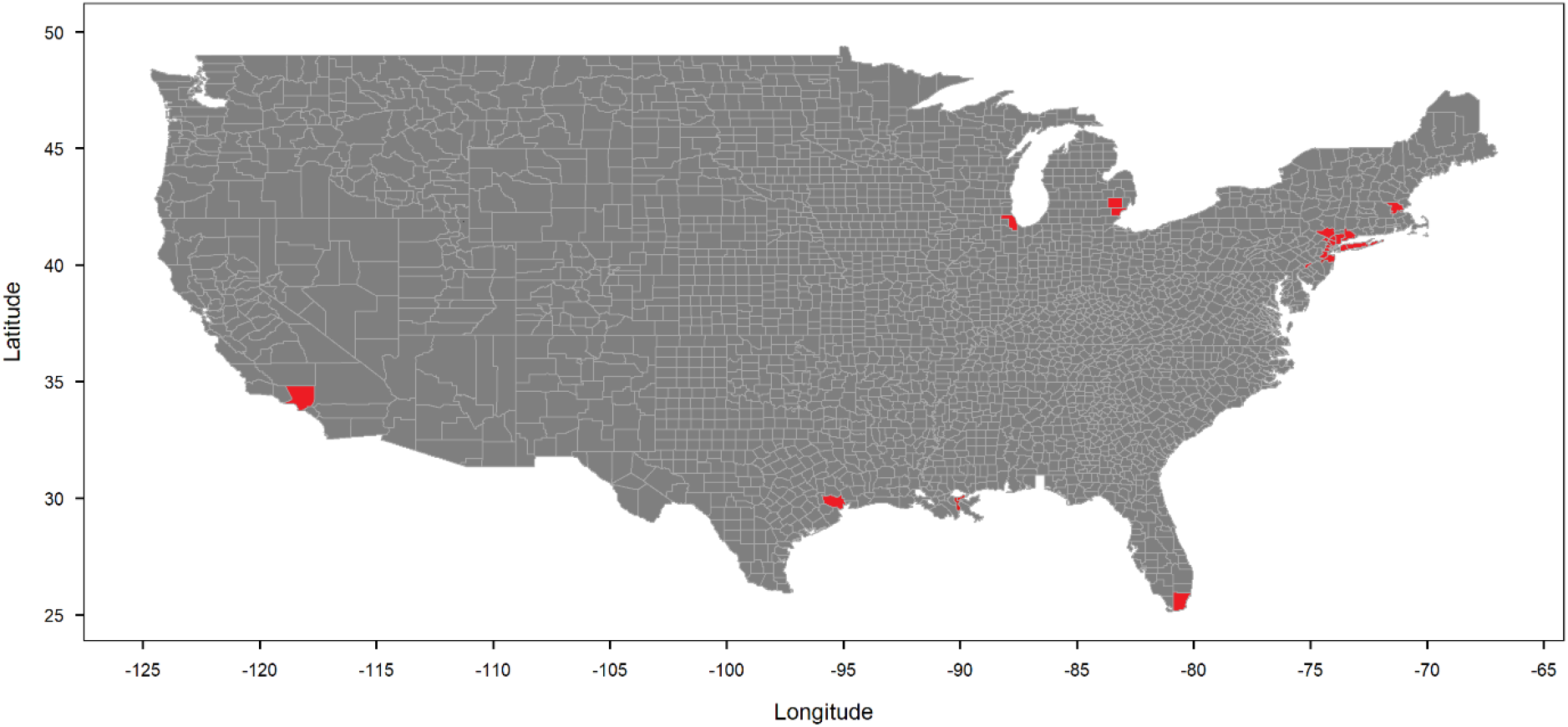
The location of the selected top 25 counties by total number of confirmed cases as of April 17.

**Fig. 3.**
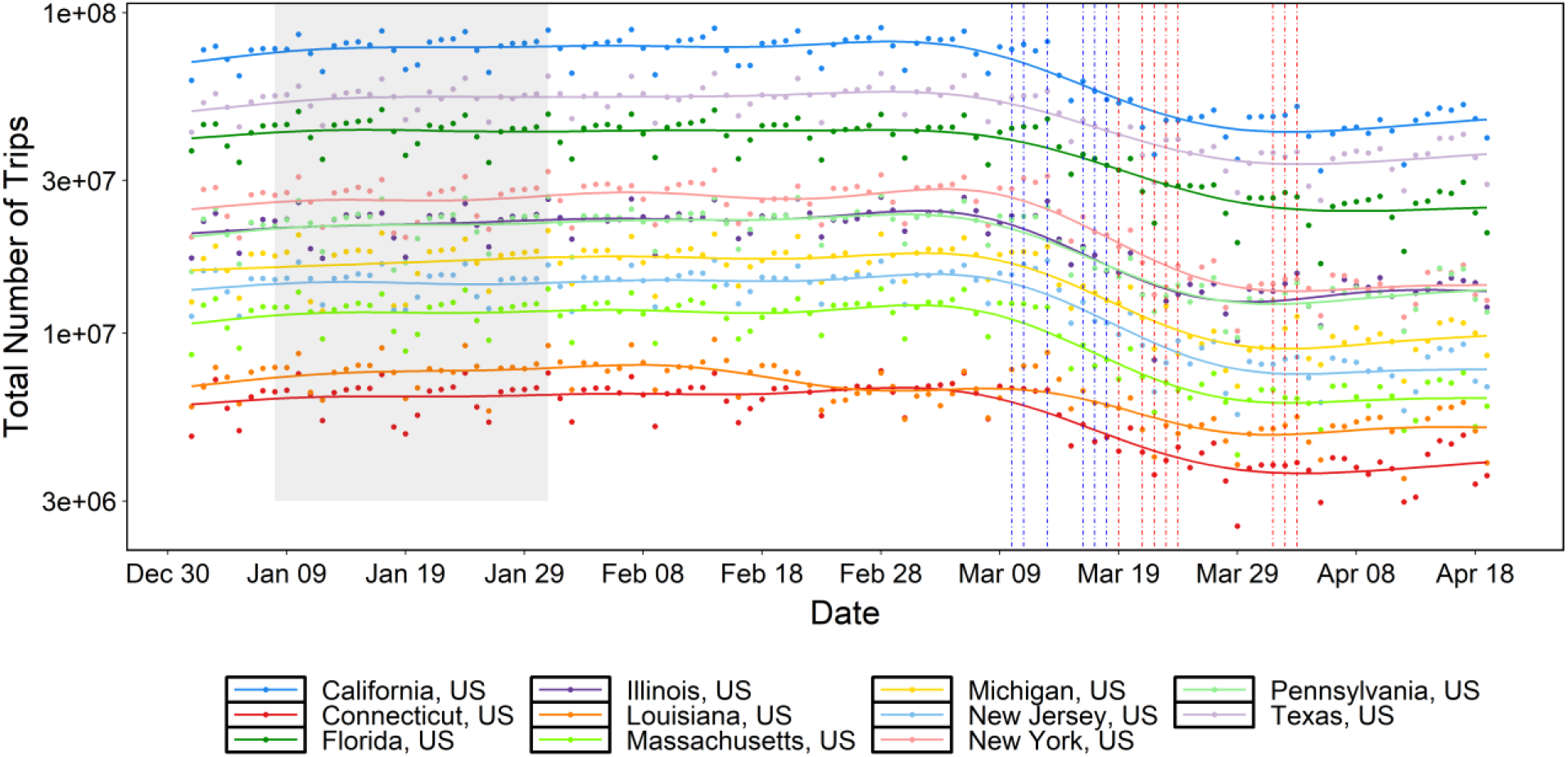
The total number of trips representing the sum of inflows, outflows, and internal trips for each state. The shaded window is the baseline period used to compute the social distancing ratio (SD). The dates of stay-at-home orders are shown as vertical dashed red and blue lines for each state and county, respectively (some dates overlap). The dots represent the raw mobility data while the plotted lines are smoothed using generalized additive model (GAM).

**Fig. 4.**
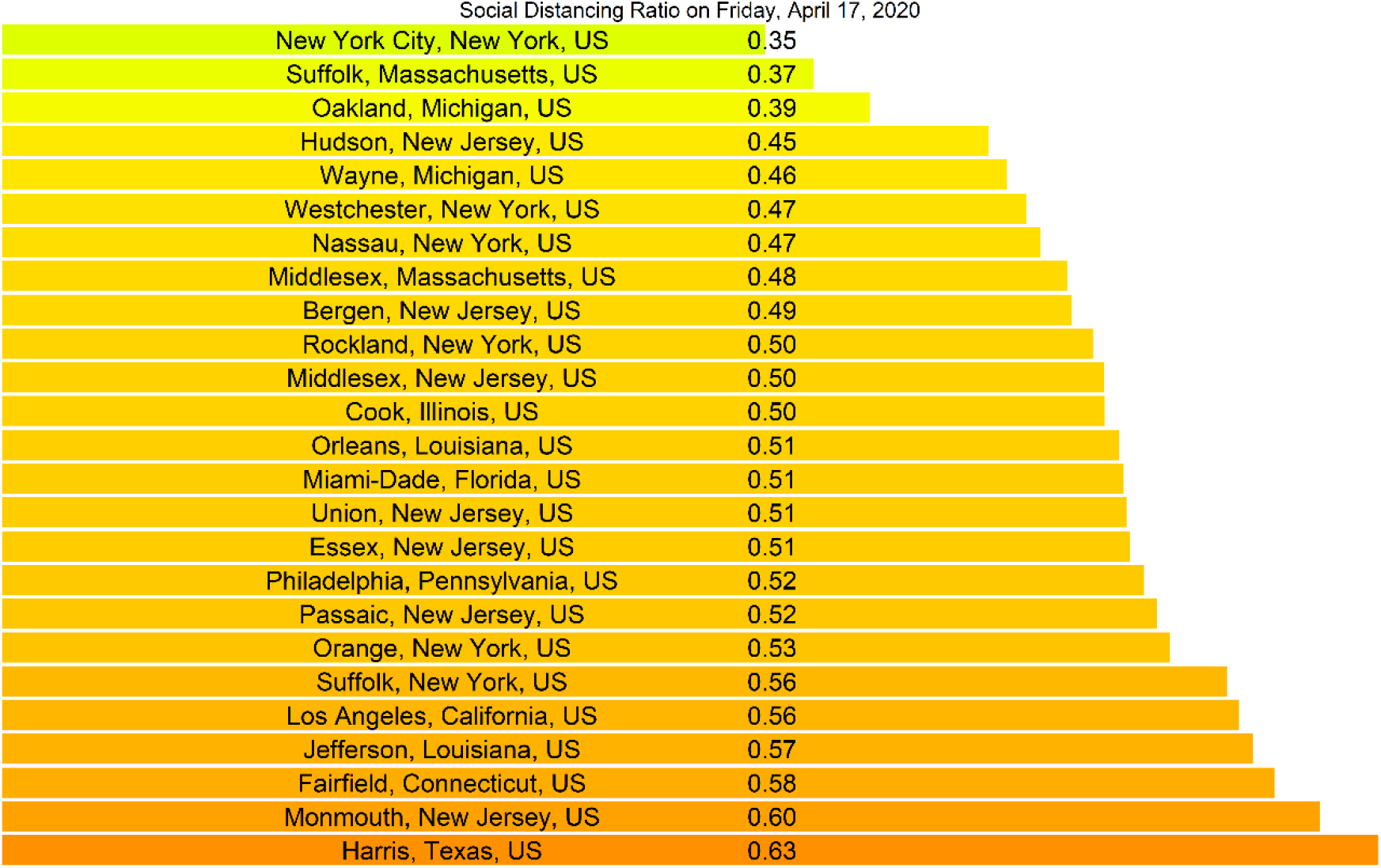
The social distancing ratio for the selected top 25 counties by total number of confirmed cases on April 17, in ascending order.

**Fig. 5.**
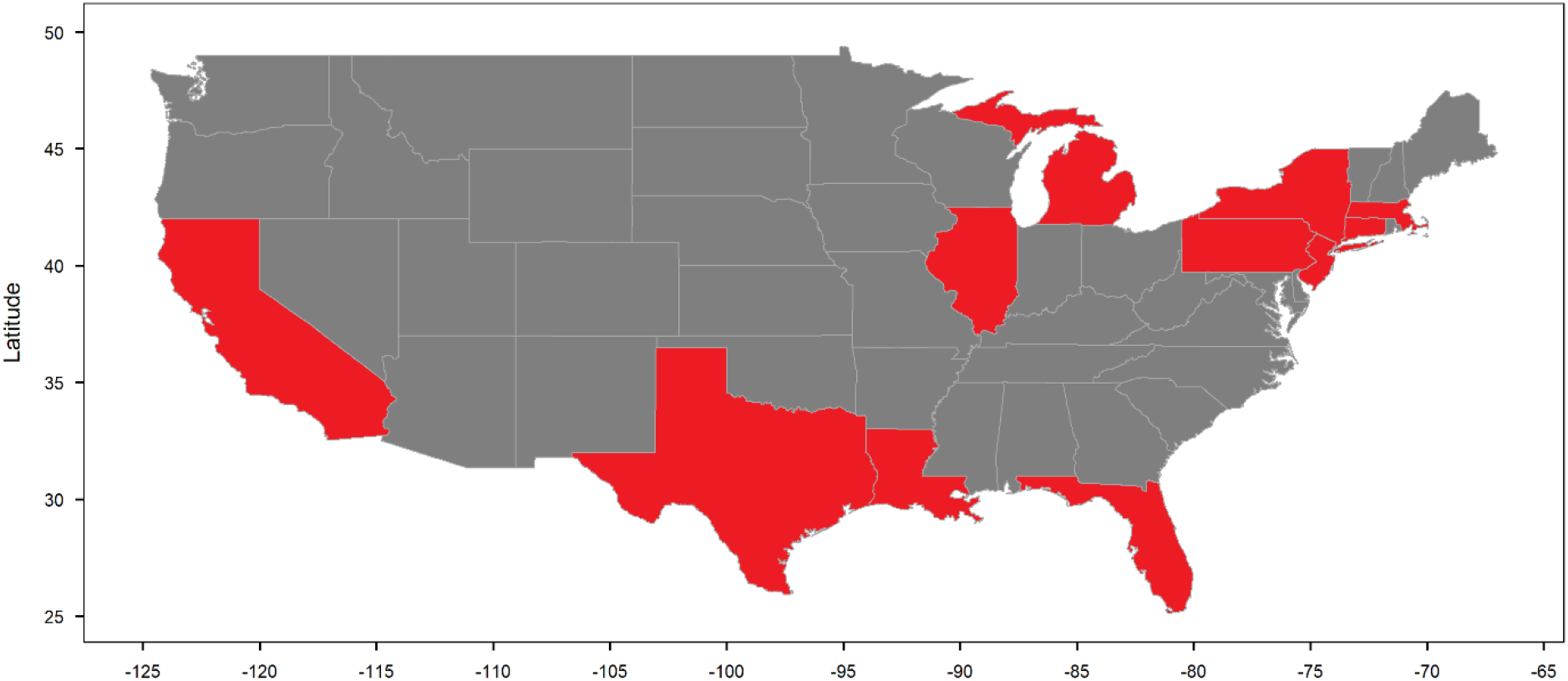
The location of the selected US states corresponding to the top 25 counties by total number of confirmed cases as of April 17.

**Fig. 6.**
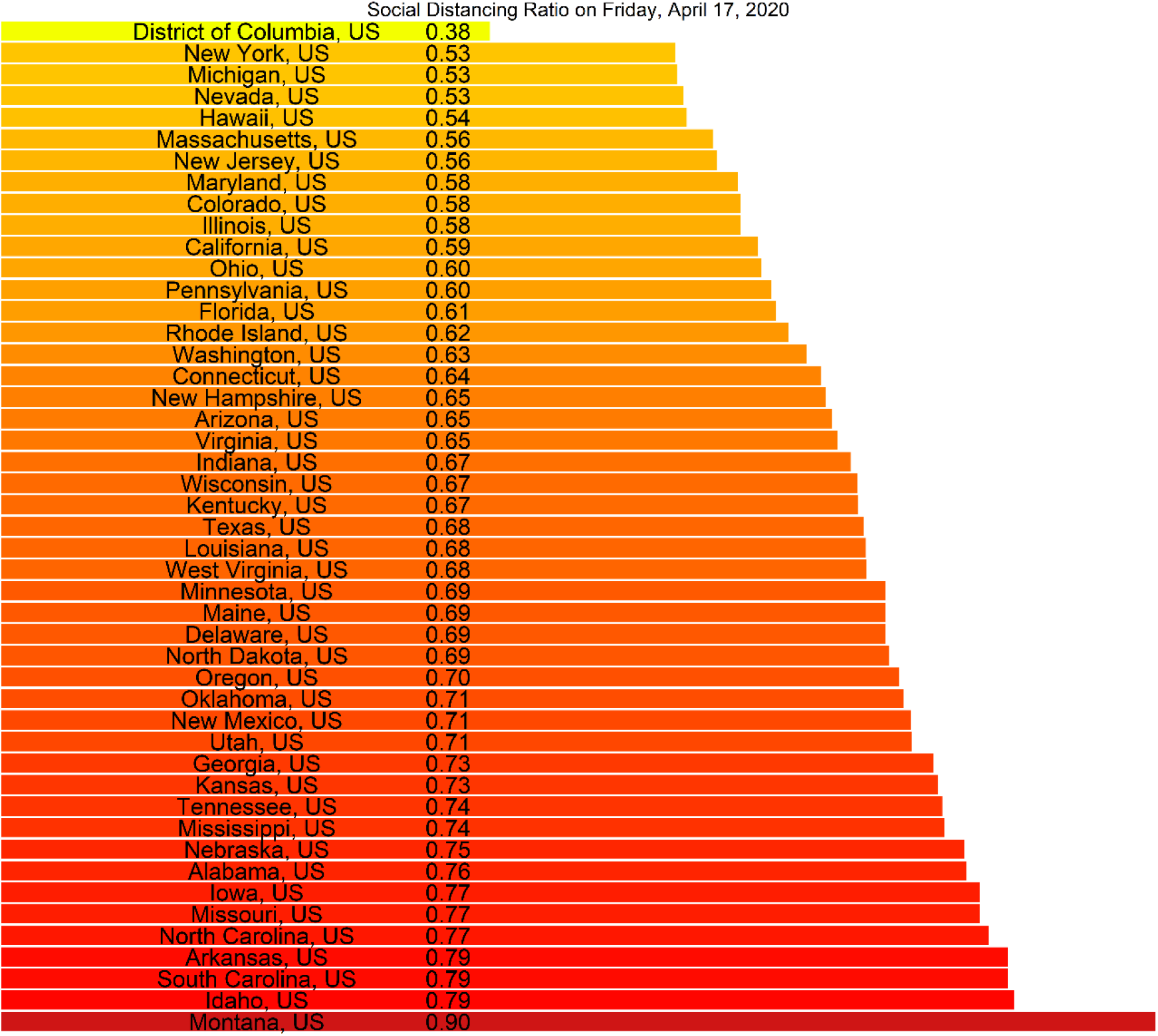
The social distancing ratio for all US states on Friday, April 17, 2020; in ascending order.

**Fig. 7.**
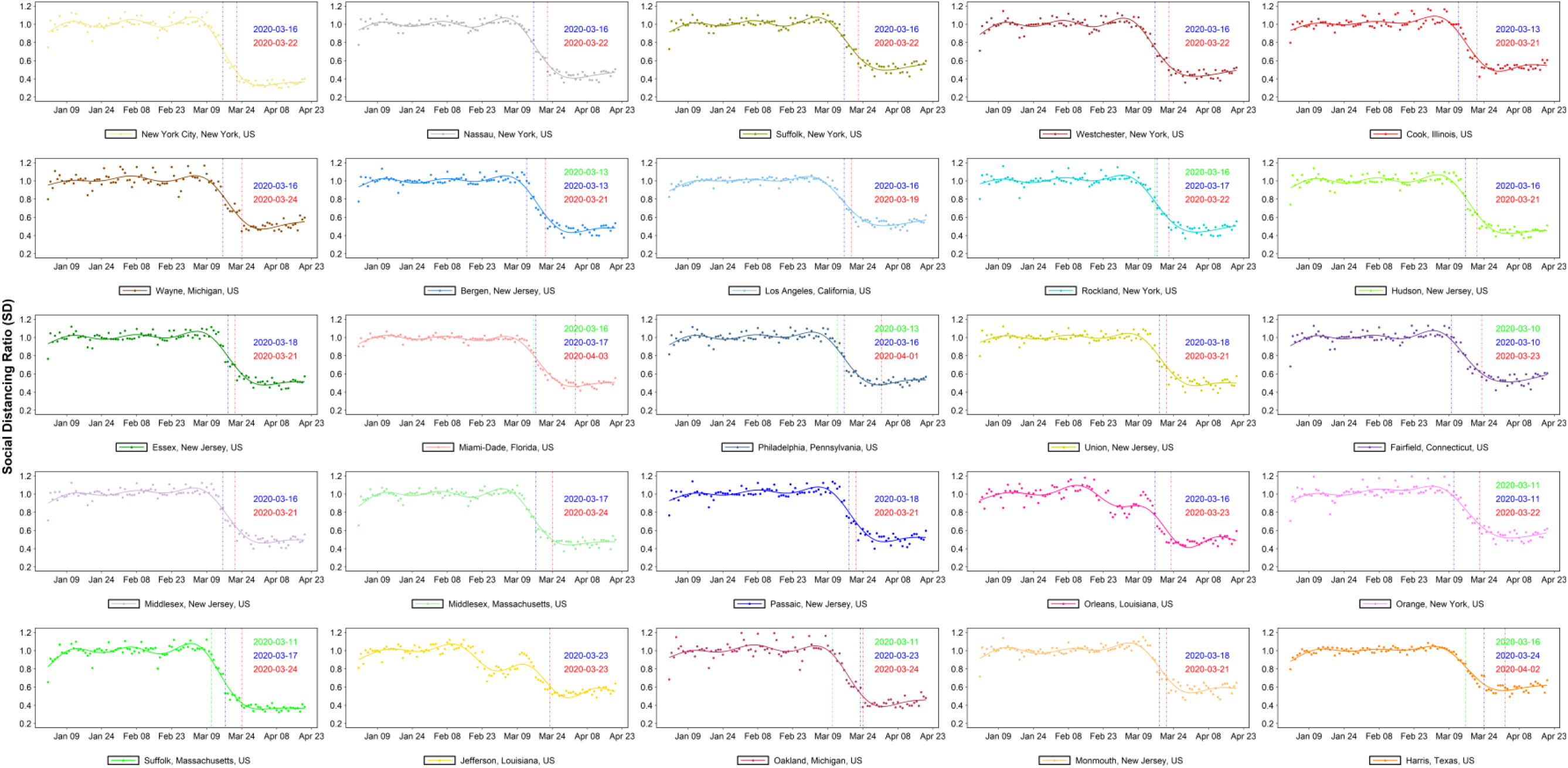
The timeseries of social distancing ratio (SD) for the selected top 25 counties. The dates of state-level stay-at-home orders are shown as vertical dashed red lines, while the school-closures and county-level social distancing orders are shown as dashed green and blue line, respectively. The dots represent the raw data while the plotted lines are smoothed using generalized additive model (GAM).

**Fig. 8.**
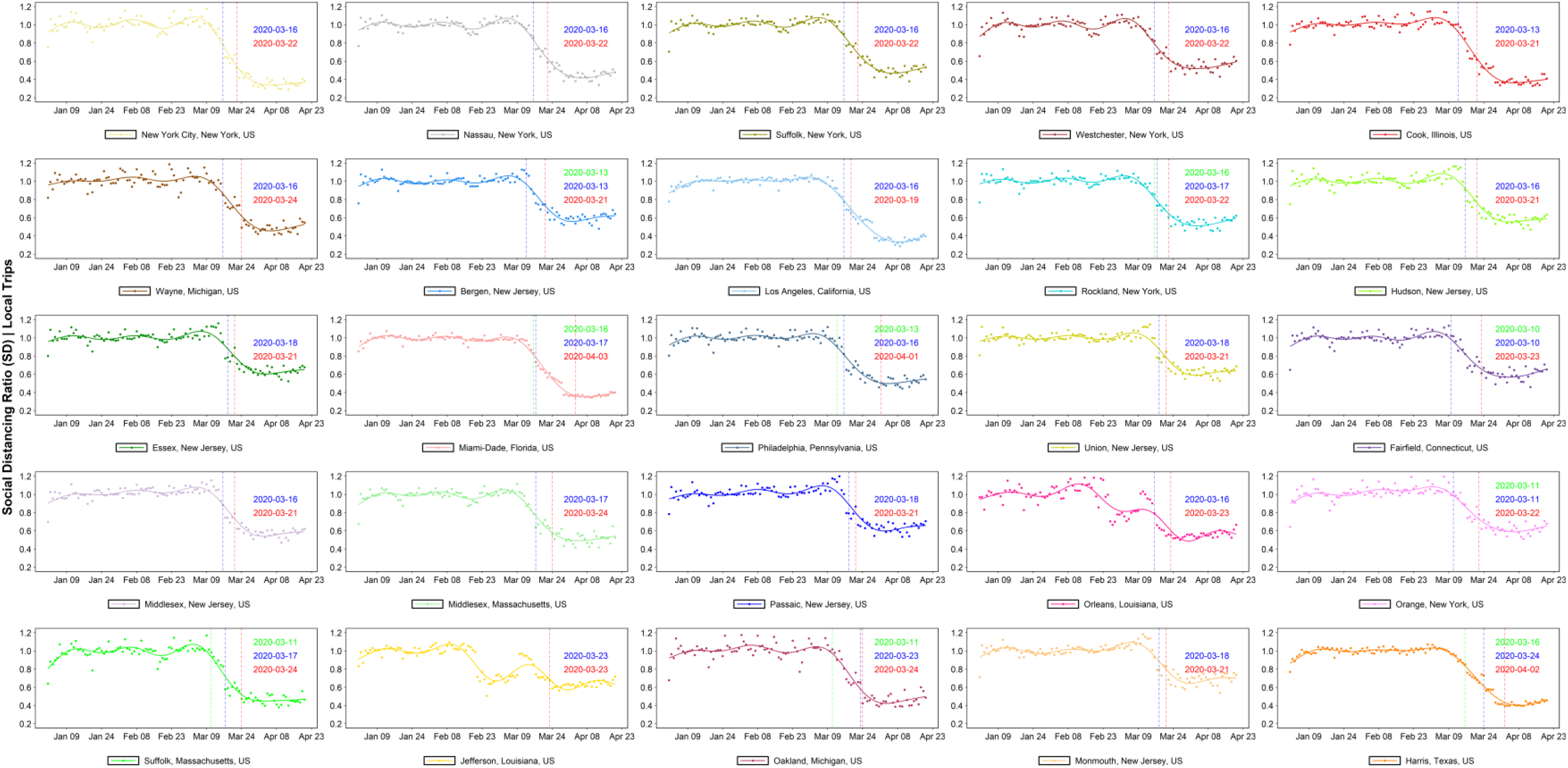
The timeseries of social distancing ratio (SD) based on internal trips (within county) only, for the selected top 25 counties. The dates of state-level stay-at-home orders are shown as vertical dashed red lines, while the school-closure and county-level social distancing orders are shown as dashed green and blue line, respectively. The dots represent the raw data while the plotted lines are smoothed using generalized additive model (GAM).

**Fig. 9.**
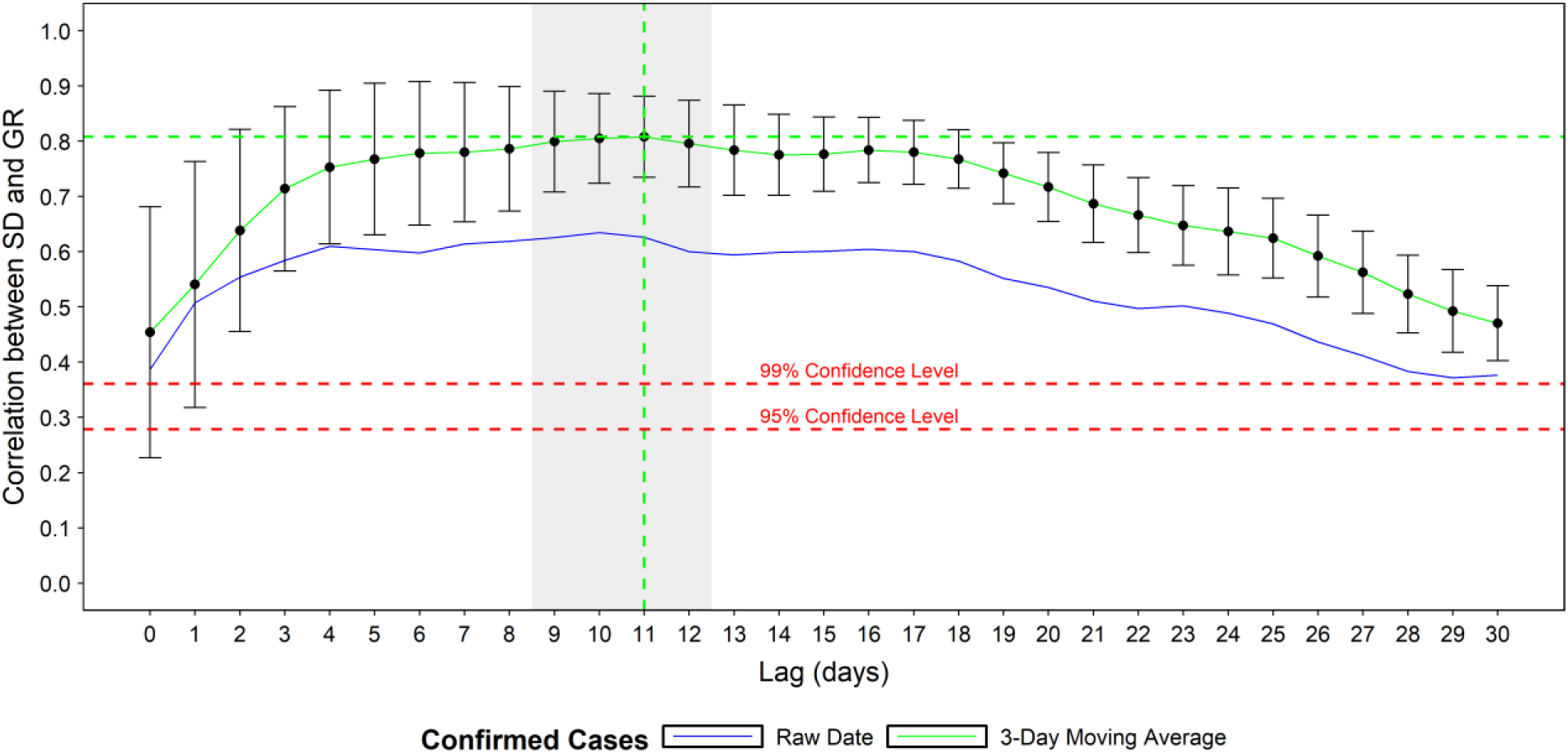
The mean and standard deviation of the state-level correlations between social distancing ratio and growth rate ratio at different lags (in days). All correlations are significant at 95% confidence level. The shaded window represents the optimal social distancing lag of nine to twelve days.

**Fig. 10.**
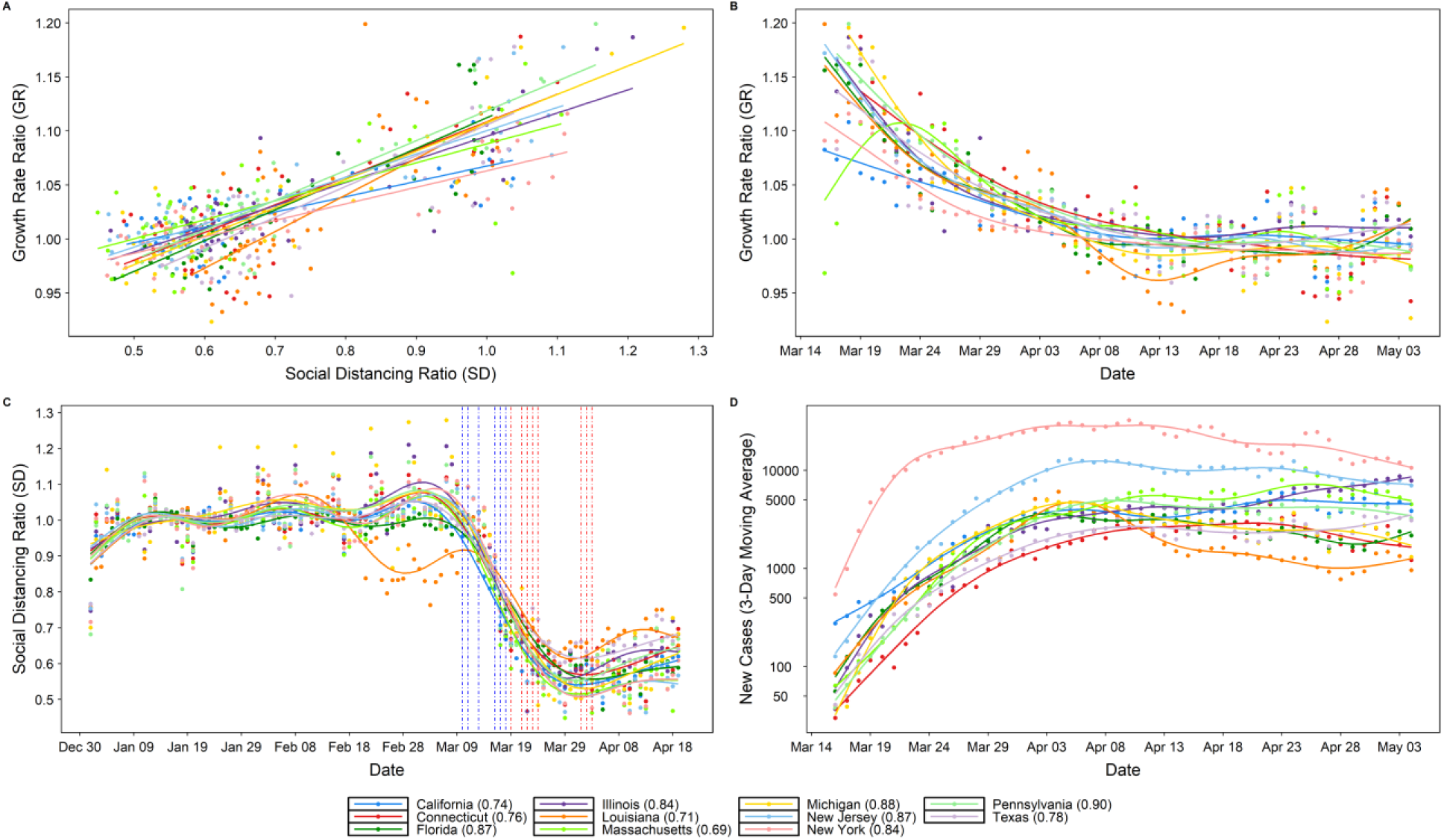
Relationship between social distancing ratio SD and growth rate ratio GR given an eleven-day lag (A), with (B) growth rate ratio GR over time (C) social distancing ratio SD over time and (D) daily confirmed cases over time. Correlations are found to be significant at a 95% confidence level. The dates of stay-at-home orders are shown as vertical dashed red and blue lines for each state and county, respectively (some overlap) in (C). The dots represent the raw data while the plotted lines are smoothed using generalized additive model (GAM).

**Table S1.**
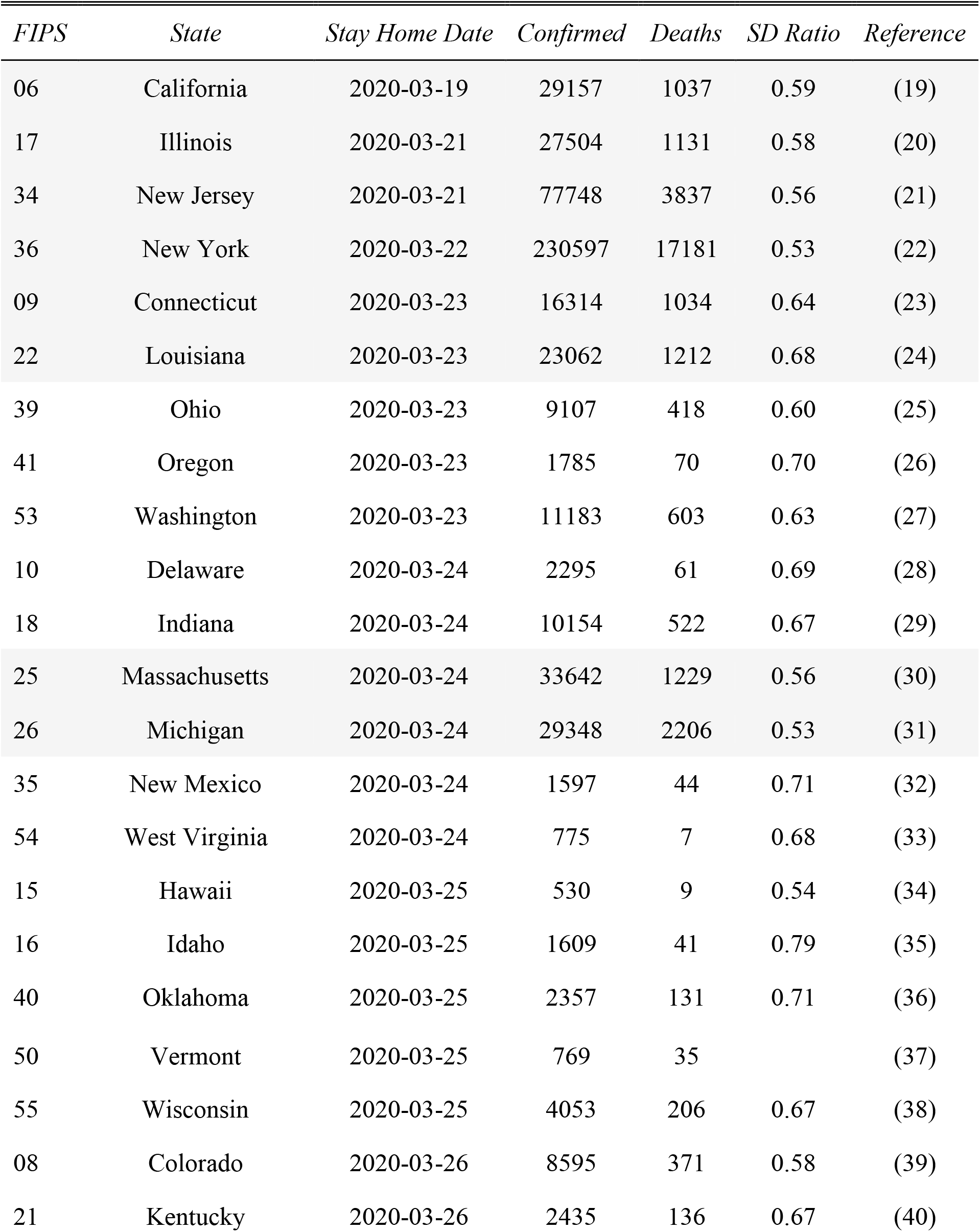
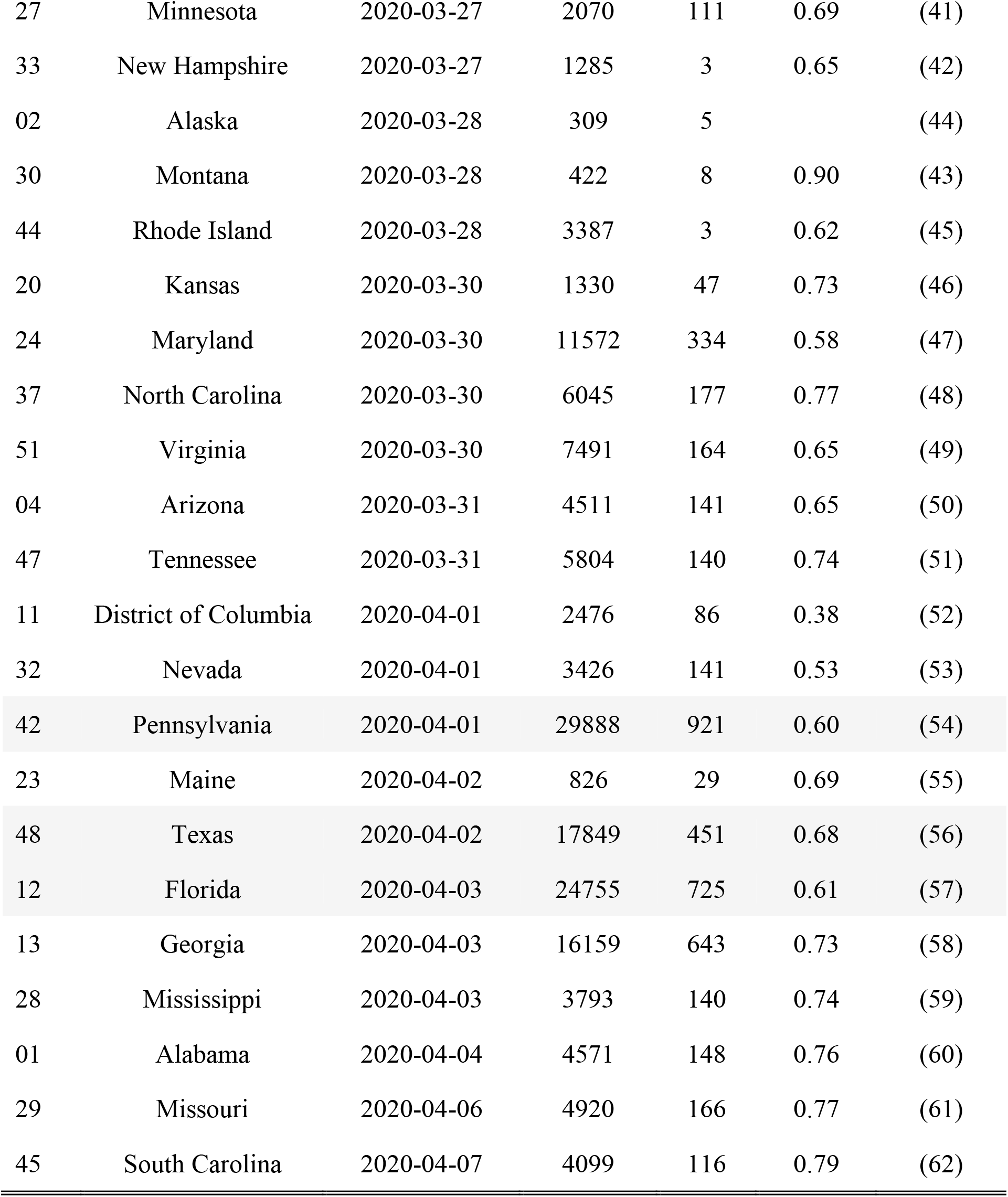
Dates and times of stay-at-home orders for each US state, and the number of confirmed cases, deaths, and social distancing ratio (SD) on April 17, 2020. The list is sorted by the stay-home order date. The 11 states highlighted are the focus of this study.

**Table S2.**
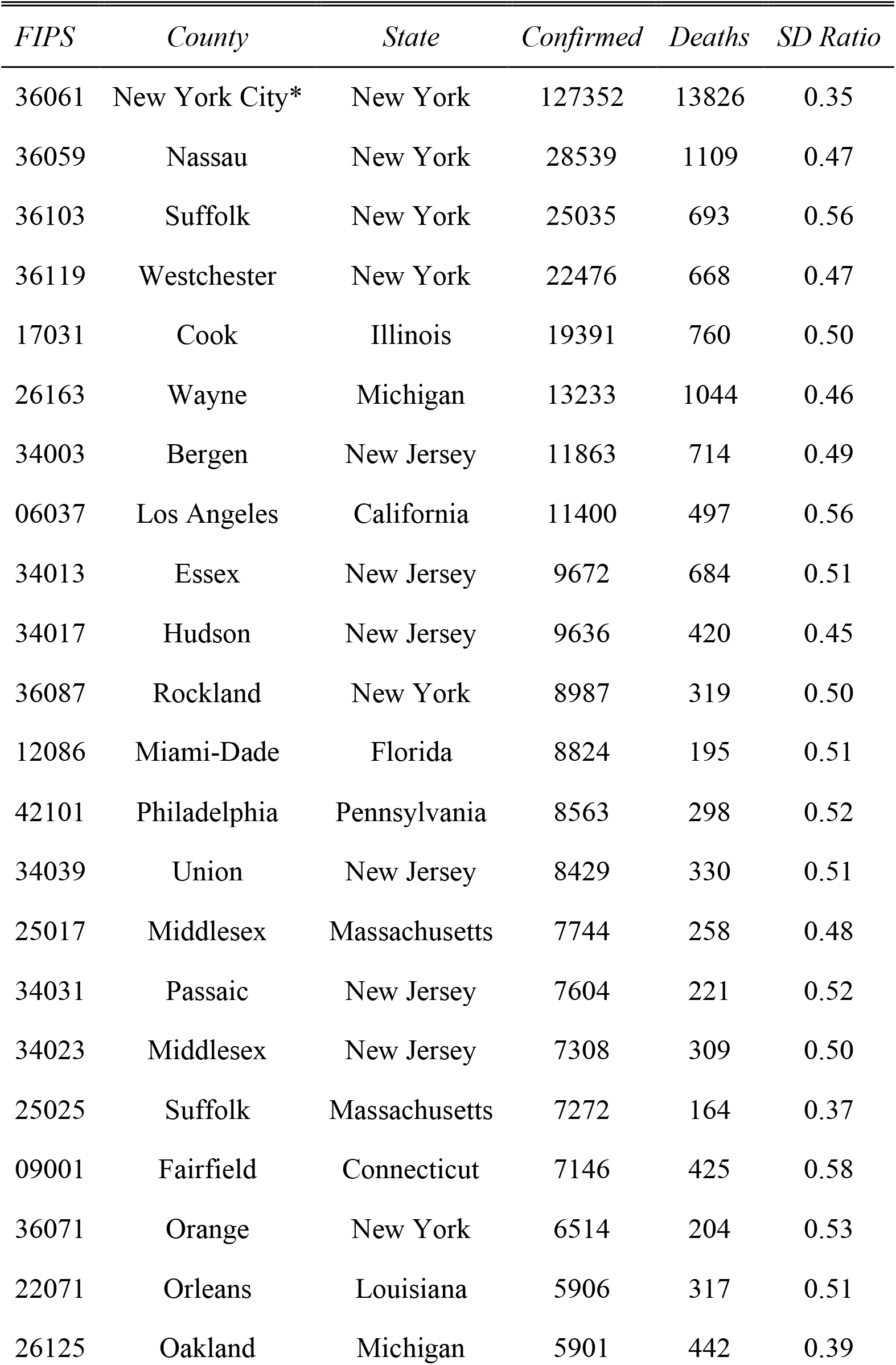
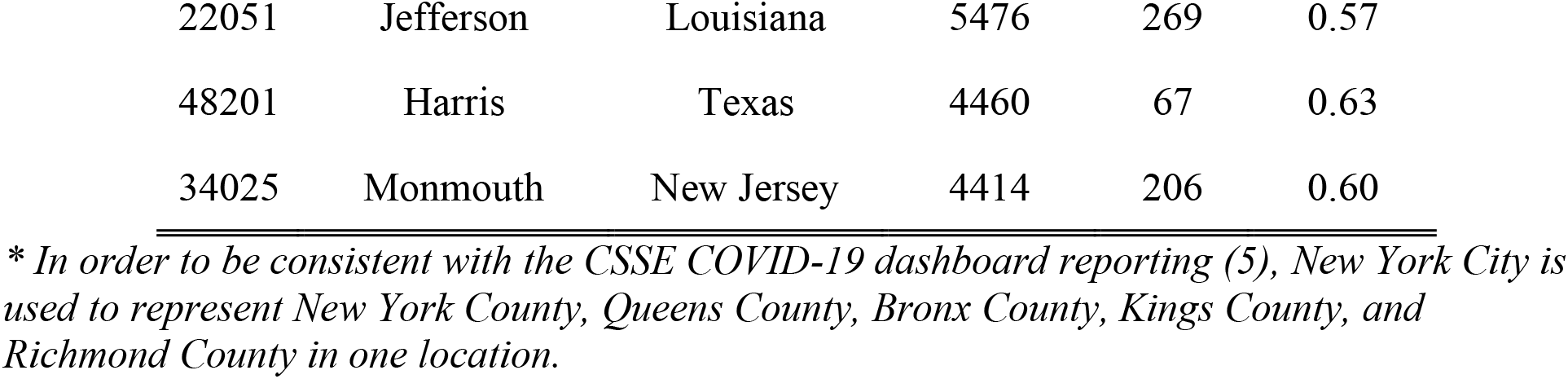
The top 25 US counties based on total cases, number of confirmed cases, deaths, and social distancing ratio (SD) on April 17, 2020.

| *FIPS* | *County* | *State* | *Confirmed* | *Deaths* | *SD Ratio* |
| --- | --- | --- | --- | --- | --- |
| 36061 | New York City* | New York | 127352 | 13826 | 0.35 |
| 36059 | Nassau | New York | 28539 | 1109 | 0.47 |
| 36103 | Suffolk | New York | 25035 | 693 | 0.56 |
| 36119 | Westchester | New York | 22476 | 668 | 0.47 |
| 17031 | Cook | Illinois | 19391 | 760 | 0.50 |
| 26163 | Wayne | Michigan | 13233 | 1044 | 0.46 |
| 34003 | Bergen | New Jersey | 11863 | 714 | 0.49 |
| 06037 | Los Angeles | California | 11400 | 497 | 0.56 |
| 34013 | Essex | New Jersey | 9672 | 684 | 0.51 |
| 34017 | Hudson | New Jersey | 9636 | 420 | 0.45 |
| 36087 | Rockland | New York | 8987 | 319 | 0.50 |
| 12086 | Miami-Dade | Florida | 8824 | 195 | 0.51 |
| 42101 | Philadelphia | Pennsylvania | 8563 | 298 | 0.52 |
| 34039 | Union | New Jersey | 8429 | 330 | 0.51 |
| 25017 | Middlesex | Massachusetts | 7744 | 258 | 0.48 |
| 34031 | Passaic | New Jersey | 7604 | 221 | 0.52 |
| 34023 | Middlesex | New Jersey | 7308 | 309 | 0.50 |
| 25025 | Suffolk | Massachusetts | 7272 | 164 | 0.37 |
| 09001 | Fairfield | Connecticut | 7146 | 425 | 0.58 |
| 36071 | Orange | New York | 6514 | 204 | 0.53 |
| 22071 | Orleans | Louisiana | 5906 | 317 | 0.51 |
| 26125 | Oakland | Michigan | 5901 | 442 | 0.39 |
| 22051 | Jefferson | Louisiana | 5476 | 269 | 0.57 |
| 48201 | Harris | Texas | 4460 | 67 | 0.63 |
| 34025 | Monmouth | New Jersey | 4414 | 206 | 0.60 |

\* In order to be consistent with the CSSE COVID-19 dashboard reporting (5), New York City is used to represent New York County, Queens County, Bronx County, Kings County, and Richmond County in one location.

**Table S3.** Dates of local directives for each the top 25 US counties, including the early actions taken, public school closures, and the corresponding state stay-at-home order dates. The list is sorted by date of public-school closure, and then FIPS.

| *FIPS* | *County* | *State* | *Early*<br>*County*<br>*Directives* | *Public*<br>*School*<br>*Change\** | *Local Actions Taken* | *State Stay-*<br>*Home Date* | *Time delta*<br>*(days),*<br>*Reference* |
| --- | --- | --- | --- | --- | --- | --- | --- |
| 36071 | Orange | New York | 3/11/2020 | 3/11/2020 | Postpones large events (C) | 3/22/2020 | 11, (63) |
| 34003 | Bergen | New Jersey | 3/13/2020 | 3/13/2020 | Public school ops change (C) | 3/21/2020 | 8, (64, 65) |
| 17031 | Cook | Illinois | 3/13/2020 | 3/13/2020 | Public school ops change (C) | 3/21/2020 | 8, (66) |
| 06037 | Los Angeles | California | 3/16/2020 | 3/16/2020 | Social distancing directives (C); LA<br>County O. of Edu. Public school ops<br>change (C) | 3/19/2020 | 3, (77) |
| 12086 | Miami-Dade | Florida | 3/16/2020 | 3/16/2020 | Social distancing directives, meal site<br>changes (C); M- DC Public Schools ops<br>change (C) | 4/3/2020 | 17, (79) |
| 22071 | Orleans | Louisiana | 3/16/2020 | 3/16/2020 | Postpones large events, restaurant ops<br>change (C); NOLA School ops change<br>(S, GEO) | 3/23/2020 | 7, (75) |
| 26163 | Wayne | Michigan | NA | 3/16/2020 | Detroit Public school ops change<br>(S, GEO) | 3/24/2020 | 8, (73) |
| 34017 | Hudson | New Jersey | 3/16/2020 | 3/16/2020 | Hoboken Public Schools ops change (C) | 3/21/2020 | 5, (68) |
| 34023 | Middlesex | New Jersey | 3/16/2020 | 3/16/2020 | Middlesex Public Schools ops change<br>(C) | 3/21/2020 | 6, (71) |
| 36059 | Nassau | New York | NA | 3/16/2020 | Public school ops change (S, GEO) | 3/22/2020 | 6, (69) |
| 36061 | New York City** | New York | NA | 3/16/2020 | Public school ops change (S, GEO) | 3/22/2020 | 6, (67) |
| 36103 | Suffolk | New York | NA | 3/16/2020 | Public school ops change (S, GEO) | 3/22/2020 | 6, (81) |
| 36119 | Westchester | New York | NA | 3/16/2020 | Public school ops change (S, GEO) | 3/22/2020 | 6, (70) |
| 42101 | Philadelphia | Pennsylvania | 3/13/2020 | 3/16/2020 | Postpones large events (C); School District of Philadelphia ops change (C) | 4/1/2020 | 16, (74) |
| 09001 | Fairfield | Connecticut | 3/10/2020 | 3/17/2020 | Postpones large events, social distancing directives; Fairfield Pub. Schools ops change (C) | 3/23/2020 | 13, (80) |
| 25017 | Middlesex | Massachusetts | NA | 3/17/2020 | Mass. Public schools' ops change (S, GEO) | 3/24/2020 | 7, (84) |
| 25025 | Suffolk | Massachusetts | 3/11/2020 | 3/17/2020 | Partial public-school ops change (C); Boston Public Schools Ops change (C) | 3/24/2020 | 13, (81) |
| 36087 | Rockland | New York | 3/16/2020 | 3/17/2020 | Social distancing, restaurant ops directive (C); Rockland County Public Schools ops change (C) | 3/22/2020 | 6, (81) |
| 34013 | Essex | New Jersey | NA | 3/18/2020 | NJ public schools' ops change (S, GEO) | 3/21/2020 | 3, (86) |
| 34025 | Monmouth | New Jersey | NA | 3/18/2020 | NJ public schools' ops change (S, GEO) | 3/21/2020 | 3, (86) |
| 34031 | Passaic | New Jersey | NA | 3/18/2020 | Passaic County Public Schools ops change (S, GEO) | 3/21/2020 | 3, (85) |
| 34039 | Union | New Jersey | NA | 3/18/2020 | NJ public schools' ops change (S, GEO) | 3/21/2020 | 3, (86) |
| 22051 | Jefferson | Louisiana | NA | 3/23/2020 | Public schools' ops change (S, GEO) | 3/23/2020 | 0, (89) |
| 26125 | Oakland | Michigan | 3/11/2020 | 3/23/2020 | Cancel large event (C); Detroit City Public Schools ops change (S, GEO) | 3/24/2020 | 13, (88) |
| 48201 | Harris | Texas | 3/16/2020 | 3/24/2020 | Houston and Harris County public schools' ops change, restaurant ops change (C); County stay home, stay safe (C) | 4/2/2020 | 17, (90) |

\* *County or State Directives*
\*\* *In order to be consistent with the CSSE COVID-19 dashboard reporting (5), New York City is used to represent New York County, Queens County, Bronx County, Kings County, and Richmond County in one location.*
*Note: C = County directive. S = State directive. GEO = Governor's Executive Order.*

## Notes

### Competing Interest Statement

The authors have declared no competing interest.

